# Risk of Myocarditis after Covid-19 mRNA Vaccination: Impact of Booster Dose and Dosing Interval

**DOI:** 10.1101/2022.07.31.22278064

**Authors:** Stéphane Le Vu, Marion Bertrand, Marie-Joëlle Jabagi, Jérémie Botton, Alain Weill, Rosemary Dray-Spira, Mahmoud Zureik

**Affiliations:** EPIPHARE Scientific Interest Group in Epidemiology of Health Products (French National Agency for the Safety of Medicines and Health Products - ANSM, French National Health Insurance - CNAM), Saint-Denis, France; Faculté de Pharmacie, Université Paris-Saclay, 92296, Châtenay-Malabry, France; University Paris-Saclay, UVSQ, University Paris-Sud, Inserm, Anti-infective evasion and pharmacoepidemiology, CESP, Montigny le Bretonneux, France

## Abstract

Covid-19 mRNA vaccines have been shown to be associated with a short-term increased risk of myocarditis, with the highest risk observed after the second dose compared to the first. The extent of the risk associated with more distant booster doses is less clear. Here, we aimed to assess the relation between dosing interval and the risk of myocarditis, for both the two-dose primary series and the third dose (first booster). Extending our previous matched case-control study, we included 4 890 cases of myocarditis aged 12 or more and 48 900 controls up to January 31, 2022. We found that the risk of myocarditis remained elevated after the booster dose and that longer intervals between each consecutive dose (including booster doses) may decrease the occurrence of vaccine-associated myocarditis.

## Main text

Covid-19 mRNA vaccines have been shown to be associated with a short-term increased risk of myocarditis, with the highest risk observed after the second dose relative to the first.^1^ The extent of the risk associated with more distant booster doses is less clear, with reports of a lower incidence relative to the second dose^2^ or an increase with sequential doses.^3^

Besides, it has been suggested that extending the timing between the first two doses could lower the risk of heart inflammation.^4^ While also accounting for an enhanced vaccine effectiveness, an optimal interval of 8 weeks between the two primary doses was recommended in some countries to mitigate the adverse events.^5^

In the context of additional booster programs to maintain protection, this raises the question of whether the time interval between all sequential doses plays a role in the risk of vaccine-associated myocarditis. Here, we aimed to assess the association between dosing interval and the risk of myocarditis for both the two-dose primary series and the third dose (first booster).

Extending our previous study,^6^ we conducted a matched case-control study within the entire French population aged 12 or older, from December, 27, 2020 to January, 31, 2022. We included all 4,890 cases of myocarditis admitted to French hospitals along with 48,900 controls of the general population matched for gender, age, and area of residency. Exposure was defined as vaccination with an mRNA vaccine 1 to 7 days prior to the index date.

Non-vaccinated subjects, and those vaccinated more than 21 days before the index date were considered to be non-exposed.

Odds ratios of myocarditis according to exposure to a vaccine injection within seven days, adjusted for recent SARS-CoV-2 infection and comorbidities were obtained using conditional logistic regression models (see Supplementary Methods). For the second and the third dose, three categories of dosing interval since the previous dose were defined using the tertiles of their respective observed distribution among controls.

The risk of myocarditis increased in the week following each of the first, second, and third doses of both the BNT162b (Pfizer-BioNTech) and mRNA-1273 (Moderna) vaccines, with odds ratios ranging from 1.7 (95% confidence interval [CI], 1.3 to 2.2) for the first dose of BNT162b to 19 (95%CI, 14 to 25) for the second dose of mRNA-1273 (Table). Odds ratios were 3.1 (95%CI, 2.3 to 4.3) after the third dose of BNT162b and 4.1 (95%CI, 2.5 to 6.6) after the third dose of mRNA-1273 vaccine.

**Table.**
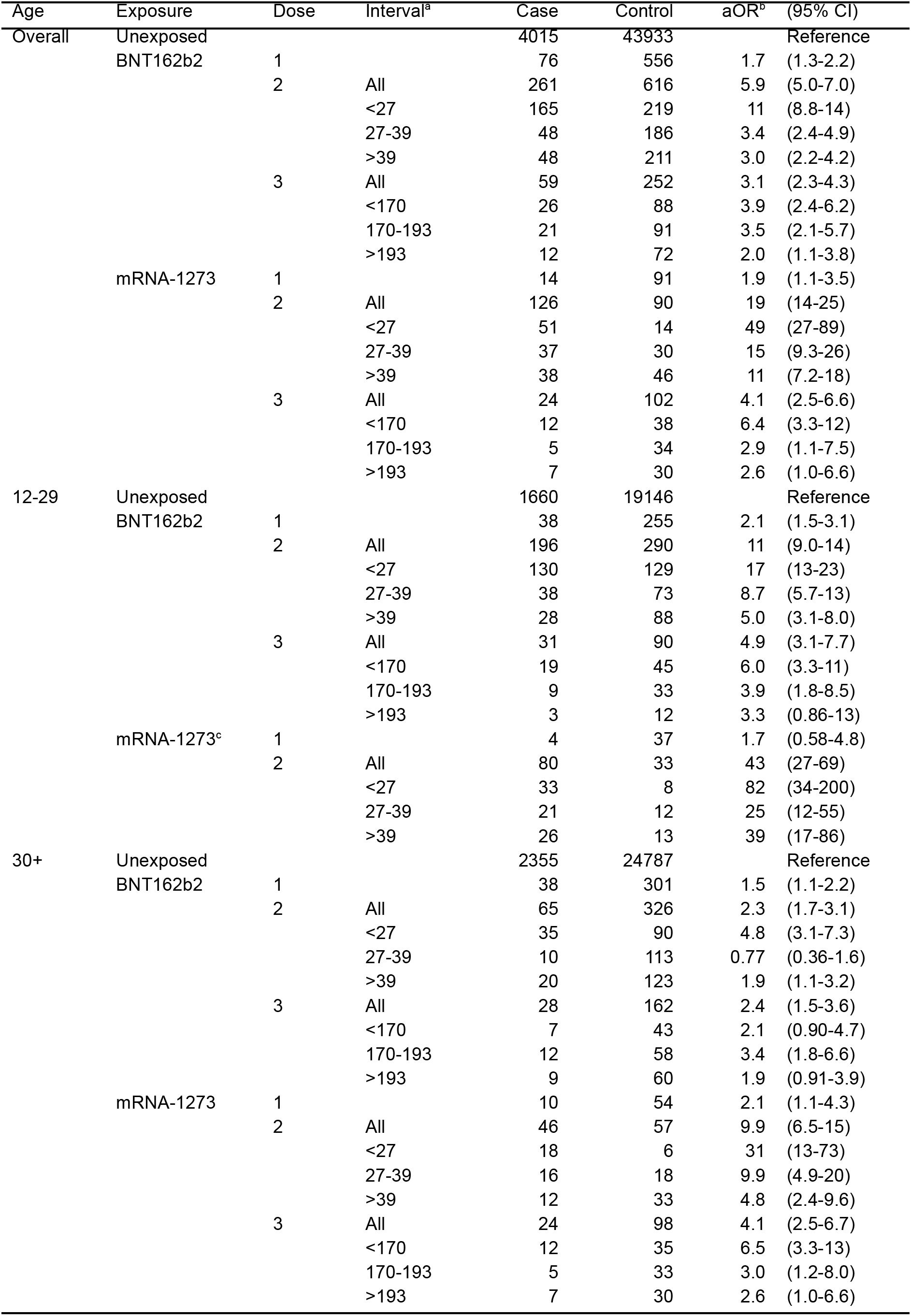
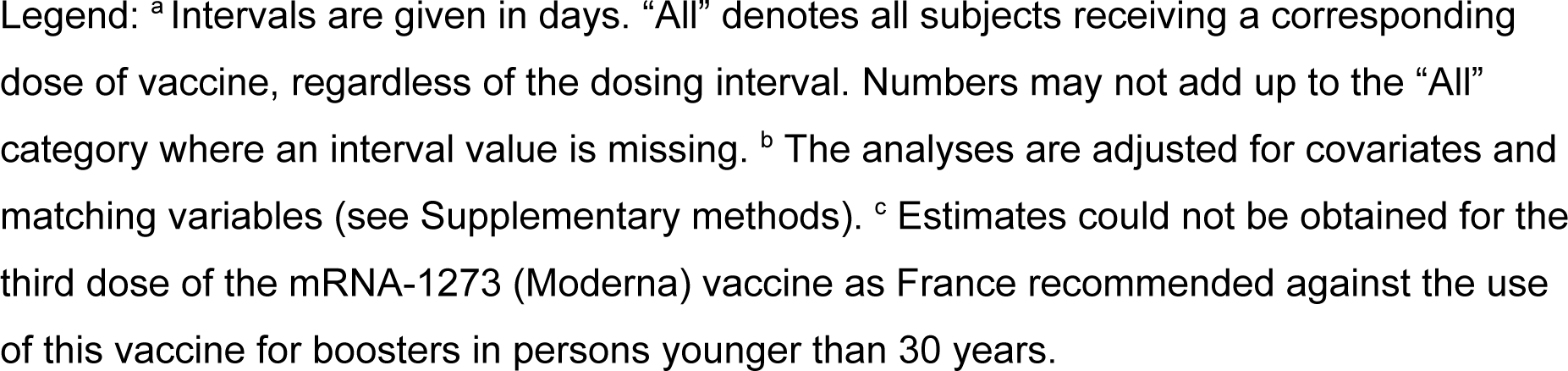
Association between myocarditis and exposure to mRNA vaccines within 7 days by age group.

Cutoffs of dosing interval tertiles were at 27 and 39 days for the second dose, and 170 and 193 days for the third dose (Figure 1a). For both vaccines, while the risk following each dose was increased regardless of dosing interval tertile, odds ratios tended to decrease as intervals grew longer (Figure 1b).

**Figure 1.**
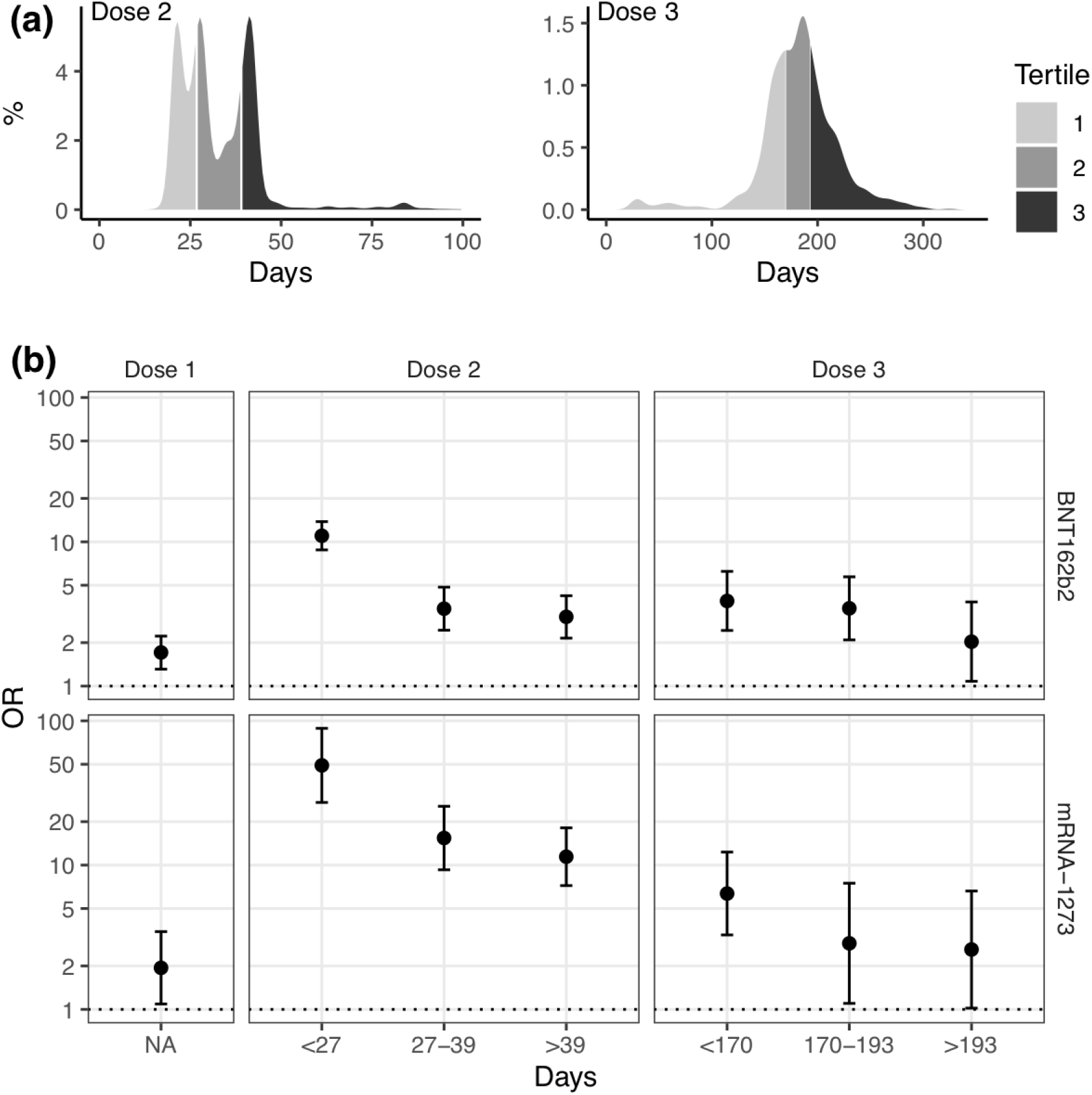
Distribution of mRNA vaccine dosing interval and odds ratio of myocarditis by interval. Legend: **(a)** Distribution of mRNA vaccine dosing interval. Dosing intervals (in days) are partitioned by tertile of delays since the previous dose (27 and 39 days between first and second doses, and 170 and 193 days between second and third doses). For each respective tertile, the median intervals were 21, 28 and 42 days (3, 4, and 6 weeks) between first and econd doses, and 156, 183, and 211 days (roughly 5, 6, and 7 months) between the second and third doses. **(b)** Odds ratio of myocarditis by dosing interval. mRNA vaccines are presented by row and doses by column. Adjusted odds ratio (OR) are represented on a logarithmic scale, with value 1 as a horizontal dotted line.

The highest risks were found for the mRNA-1273 administered within the lowest interval tertile (odds ratios of 49 [95%CI, 27 to 89] for the second dose and 6.4 [95%CI, 3.3 to 12] for the third dose). This pattern was mostly observed when stratifying by age (Table).

In this study, the risk of myocarditis remained elevated after the booster dose, albeit at a lower level than following the second dose. This result observed in all age categories from age 12, extends prior observations for young adults.^2^ Our findings bring new evidence suggesting that longer intervals between each consecutive dose (including booster doses) may decrease the occurrence of vaccine-associated myocarditis. These findings have important public health implications and suggest that, in addition to the need of maintaining immunity, the occurrence of heart inflammation should be accounted for when defining the schedules for additional booster dose vaccination.^5^

## Data Availability

According to data protection and the French regulation, the authors cannot publicly release the data from the French national health data system (SNDS). However, any person or structure, public or private, for-profit or non-profit, is able to access SNDS data upon authorization from the French Data Protection Office (CNIL Commission Nationale de l'Informatique et des Libertes) to carry out a study, a research, or an evaluation of public interest (https://www.snds.gouv.fr/SNDS/Processus-d-acces-aux-donnees and https://www.indsante.fr/).

## Author Contributions

S.L.V., M.B., A.W., R.D.S. and M.Z. conceived the study. A.W., R.D.S. and M.Z. supervised the project. M.B. carried out the clinical data collection and data curation. S.L.V. and M.B. designed and performed the statistical analyses with M.J.J. and J.B. providing input. S.L.V. wrote the first draft of the manuscript. All authors interpreted the results, provided critical revision of the manuscript and approved its final version for submission.

## Additional Contributions

We thank Bérangère Baricault and Jérôme Drouin (EPIPHARE, Saint-Denis, France) for their technical support related to data management.

## Supplementary information

### Supplementary methods

Extending our previous study^1^, we conducted a matched case-control study within the entire French population aged 12 or older, from the beginning of the Covid-19 vaccination campaign in December, 27, 2020 to January, 31, 2022.

The study was based on data of the National Health Data System (SNDS) which covers more than 99% of the French population (67 million inhabitants).^2,3^ Data on hospital admission were obtained from the French hospital discharge database (PMSI) and linked at the individual level with the nationwide databases for Covid-19 vaccination (VAC-SI) and testing (SI-DEP).

Our research group (EPI-PHARE) has permanent regulatory access to the data from the SNDS. This permanent access is given according to the French Decree No. 2016-1871 of December 26, 2016 relating to the processing of personal data called “National Health Data System” and French law articles Art. R. 1461-13 and 14. This study was declared prior to initiation on the EPI-PHARE registry of studies requiring the use of the SNDS (n° EP-0311).

Cases corresponded to all patients admitted to French hospitals with a diagnosis of myocarditis in the study period. Diagnoses at hospital were typically based on presenting symptoms, electrocardiography, echocardiography and cardiac, magnetic resonance imaging.^4,5^ We used the codes for myocarditis (I40.x, I41.x, and I51.4) of the International Classification of diseases, 10th revision (ICD-10) for detection.

Each case was matched at the date of his/her hospital admission for myocarditis (index date) to 10 control individuals. Controls were selected from among the general population by simple random sampling without replacement within each stratum of age, gender and area of residence (matching criteria), with constraint of not being diagnosed with myocarditis and being alive at the index date.

Exposure was defined as vaccination with an mRNA vaccine 1 to 7 days prior to the index date, considering the first, second and third dose separately. Non-vaccinated subjects, and those vaccinated more than 21 days before the index date were considered to be non-exposed. Of note, France suspended the use of the mRNA-1273 (Moderna) vaccine for all population groups on October, 15, 2021, and made it available again for boosters and primary series on November, 8, 2021 – but at half-dose and only for those over 30 years old.^6^

In addition to the matching variables (age, gender, and area of residence) and the initial covariates^1^ potentially associated with a risk of myocarditis or vaccine exposure (history of myocarditis in last 5 years, SARS-CoV-2 infection within a month, and socio-economic level), we included a set of covariates in the analyses. These binary variables reflected a prior history (within 5 years unless otherwise mentioned) of the following conditions: sarcoidosis, opioid use disorder, coronary artery disease and myocardial infarction, heart failure, antihypertensive drug use, arrhythmia, diabetes, autoimmune disease, respiratory disease, active cancer, and antibiotic use (within a month).^2^

We used conditional logistic regression models to estimate the odds ratios of myocarditis associated with exposure to mRNA vaccination within a week, adjusted for covariates and matching variables. The analyses were conducted according to the ranking of vaccine dose (first, second or third dose) across the entire study group and separately for males and females and by age brackets (12–29, 30 years or more). Furthermore, vaccine exposure was categorized by the time interval between two consecutive doses. For the second and the third dose, three categories of dosing interval were defined by the 33^rd^ and 66^th^ percentiles of all observed intervals since previous dose among the control population, resulting in three tertiles of intervals.

Analyses were performed using R version 4.1.2 (R Foundation for Statistical Computing).

